# The spatiotemporal estimation of the dynamic risk and the international transmission of 2019 Novel Coronavirus (COVID-19) outbreak: A global perspective

**DOI:** 10.1101/2020.02.29.20029413

**Authors:** Yuan-Chien Lin, Wan-Ju Chi, Yu-Ting Lin, Chun-Yeh Lai

## Abstract

An ongoing novel coronavirus SARS-CoV-2 pneumonia infection outbreak called COVID-19 started in Wuhan, Hubei Province, China, in December 2019. It both spread rapidly to all provinces in China and started spreading around the world quickly through international human movement from January 2020. Currently, the spatiotemporal epidemic transmission patterns, prediction models, and possible risk analysis for the future are insufficient for COVID-19 but we urgently need relevant information, particularly from the global perspective.

We have developed a novel two-stage simulation model to simulate the spatiotemporal changes in the number of COVID-19 cases and estimate the future worldwide risk. Based on the connectivity of countries to China and the country’s medical and epidemic prevention capabilities, different scenarios are generated to analyze the possible transmission throughout the world and use this information to evaluate each country’s vulnerability to and the dynamic risk of COVID-19.

Countries’ vulnerability to the COVID-19 outbreak from China is calculated for 63 countries around the world. Taiwan, South Korea, Hong Kong, and Japan are the most vulnerable areas. The relationship between each country’s vulnerability and days before the first imported case occurred shows a very high exponential decrease. The cumulative number of cases in each country also has a linear relationship with vulnerability, which can compare and quantify the initial epidemic prevention capabilities to various countries’ management strategies. In total, 1,000 simulation results of future cases around the world are generated for the spatiotemporal risk assessment. According to the simulation results of this study, if there is no specific medicine for it, it will likely form a global pandemic. This method can be used as a preliminary risk assessment of the spatiotemporal spread for a new global epidemic. * Note: This study was completed on February 15, 2020.

## Introduction

In December 2019, a novel coronavirus SARS-CoV-2 infection pneumonia outbreak COVID-19 (2019-nCoV) started in Wuhan, Hubei Province, China (Chan et al., 2020; Li et al., 2020; Munster, Koopmans, van Doremalen, van Riel, & de Wit, 2020). It spread rapidly to all provinces throughout China and started spreading around the world quickly through international human movement from January 2020 (Li et al., 2020; WHO, 2020). Thus far (February 2020), the epidemic has not decelerated but has started breaking out internationally. For example, cluster outbreaks have occurred in Singapore, Japan, and on international cruise ships (e.g. the Diamond Princess). With the exception of China, countries are at the most critical stage to avoid outbreaks of domestic cluster infections.

Many studies or reports have started revealing that SARS-CoV-2 is very infectious (Li et al., 2020; Wu, Leung, & Leung, 2020) Many clinical cases are asymptomatic or mildly diagnosed patients (Liu, Liao, Chang, Chou, & Lin, 2020), which greatly increases the potential for transmission and makes epidemic prevention very difficult. Therefore, it will likely evolve into a global pandemic of epidemic disease in the future.

Many countries have started implementing measures to prevent the epidemic spreading in their countries, including stopping flights from China, quarantining at airports, suspending visas for Chinese citizens, issuing travel warnings to China and advising citizens not to visit China unless necessary, and implementing home quarantine or stopping the entry and transfer of foreign tourists who have traveled to China within 14 days to prevent imported cases after overseas traveling. However, since many countries have very close business or human relationships with China, it is impossible to avoid imported cases. Therefore, to understand the initial transmission of novel infectious diseases, we must first understand each country’s connectivity to the country of origin, which is the first step for countries around the world to prevent the first stage of overseas imported cases.

Since the outbreak, we all want to know how COVID-19 will spread from Wuhan all across China and further from China to all around the world in terms of both time and space. A previous study made preliminary estimations and predictions through a typical infectious disease model called a susceptible–exposed–infectious–recovered (SEIR) model (Wu et al., 2020); however, as it is a new infectious disease, it is difficult for us to understand how the epidemic will develop in the future such as the parameters when the peak is reached. Unknown infectious diseases have made it very difficult to adjust and calibrate the parameters of various prediction models like the SEIR model, making the model predictions very uncertain and restrictive. The totally different pattern of transmission means that all spatiotemporal models or patterns from other coronaviruses such as the 2003 severe acute respiratory syndrome (SARS) outbreak and the Middle East respiratory syndrome (MERS) are less suitable for use with COVID-19, even if they have highly similar genome sequence identity (Lu et al., 2020; Qiao, 2020). In addition, since the global epidemic is just beginning, there is insufficient data to implement proper supervised learning models in machine learning methods or even statistical models.

Currently, the spatiotemporal epidemic transmission patterns, prediction models, and possible risk analyses for the future at the global-scale remain insufficient for COVID-19 but we urgently need relevant information. This is an important and critical time for global early-stage epidemic control and public health issues. It is difficult for us to predict future case changes in various countries, particularly the first stage of overseas imported cases from China and the second stage of local transmission cases. Under this background of high uncertainty around COVID-19, our study aims at global COVID-19 risk analysis from a data analysis perspective.

Understanding the connectivity with the country of origin for infectious diseases, i.e. China, is a useful way for countries to quantitatively evaluate the risk of imported cases. Higher connectivity is indicative of a higher risk of importation. In addition, for the analysis of worldwide countries other than China, we can use the changes in the number of cases that have been widespread in China’s provinces to estimate the number of cases worldwide. In other words, except for Hubei Province, where Wuhan is located, the epidemic pattern of China’s provinces precedes that of countries around the world by about 0.5–1 months. Thus, this can be used as a dynamic reference basis for case prediction, simulation, and risk analysis in various countries (Figure 1).

**Figure 1.**
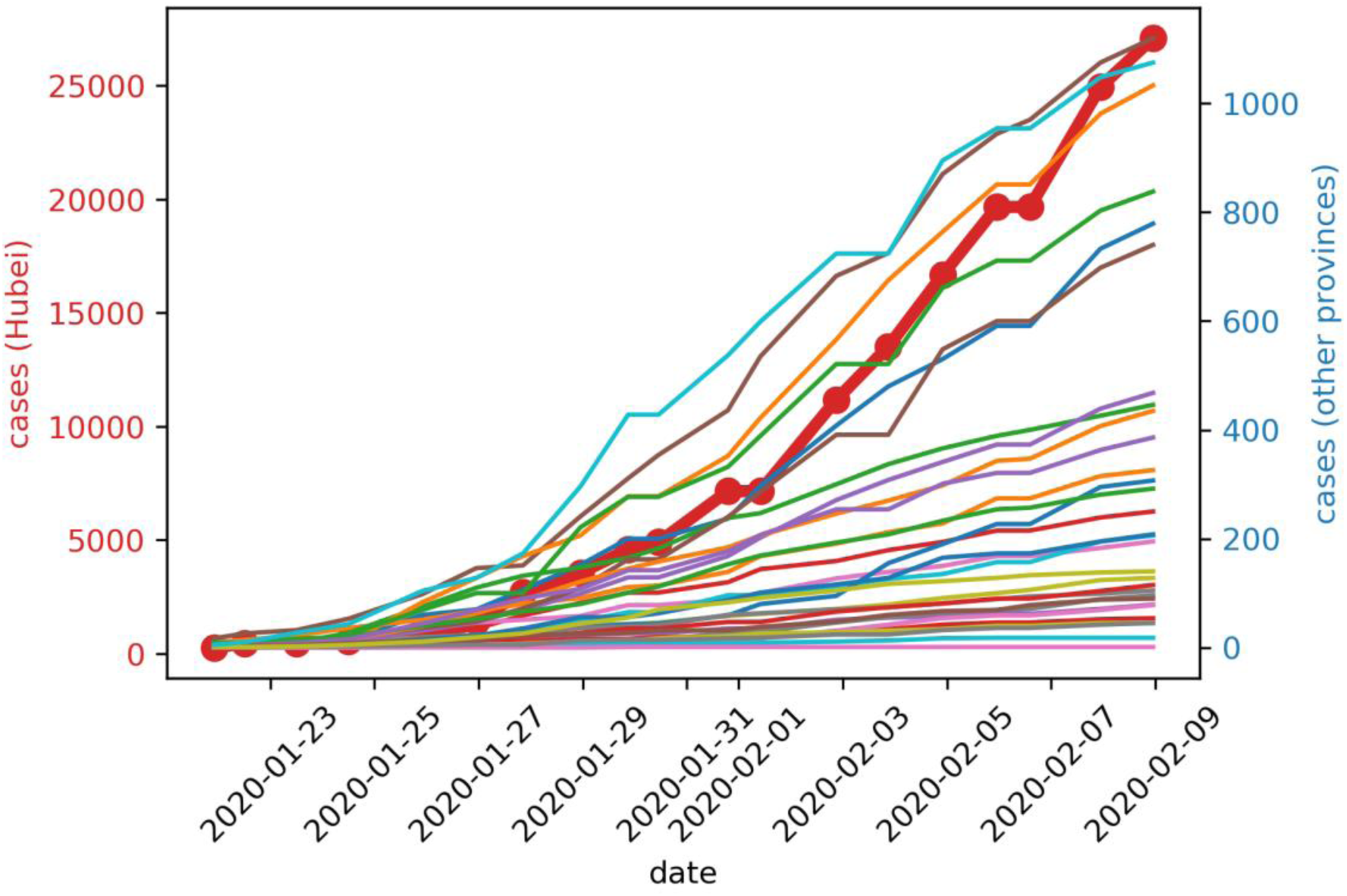
The cumulated confirmed case numbers of all provinces in China for 2020/1/21–2020/2/9

In this study, we developed a novel two-stage simulation model to simulate the spatiotemporal changes in the number of COVID-19 cases and estimate the future worldwide risk based on a data-driven approach. Based on each country’s connectivity to China, medical and epidemic prevention capabilities, and future conditions, different scenarios are generated with which to analyze the potential transmission around the world. Meanwhile, from a risk management perspective, understanding risk and proper risk management is a very important part of preventing the spread of infectious diseases in various countries. According to the definition of the Intergovernmental Panel on Climate Change (IPCC) at the United Nations, risk can be considered a function of exposure, hazard, and vulnerability (IPCC, 2014). Therefore, to assess the risk of COVID-19 from China, we defined and calculated the countries’ exposure and hazard of and vulnerability to COVID-19 around the world and generated comprehensive spatiotemporal dynamic risk maps over time.

## Methods

### a. Data

The original laboratory-confirmed cases data is collected from the WHO situation reports of COVID-19 (WHO, 2020) and the Center for Systems Science and Engineering (CSSE) at Johns Hopkins University (JHUCSSE, 2020) for 2020/1/21–2020/2/12. To define connectivity between countries, international mobility data is calculated from flight route numbers between airports. If more direct routes are found between two airports, this represents higher connectivity. The original route data is accessible from Openflight.org, which is an open-source project to collect and visualize flight information. The world’s Health Care Index is collected from Numbeo, which is the world’s largest database of user-contributed data about cities and countries worldwide. Population density data were collected from the World Bank.

### b. Two-stage simulation modeling framework

#### First stage: Imported cases

Assuming that all countries are fully committed to epidemic prevention and well-prepared for epidemic prevention but that there are no specific medicines or vaccines, it is assumed that COVID-19 is not affected by climatic factors (such as temperature). The changes in the case number for each country can be divided into two major stages. The first stage is mainly based on imported cases of immigration from abroad or sporadic cases of local traceable infection sources. The time that the first case happens in each country and the daily number of subsequent cases that continue to occur are based on the country’s Health Care Index and connectivity to the country of outbreak. Here, the country’s vulnerability to outbreak countries for coronavirus is defined as:

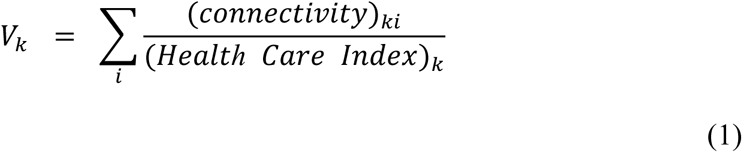

where V is the vulnerability of country *k, i* is the outbreak countries, the connectivity is defined as the total number of flight routes between country *k* and country *i*, the outbreak countries. The final vulnerability value is standardized to 0–1.

The higher the Health Care Index, the better the resistance against outbreaks from outbreak countries so we have less vulnerability. Meanwhile, higher connectivity or transmissibility indicates that a country has more interactions and higher human mobility with outbreak countries, which makes the disease more easily transmitted to this country, so it has a higher vulnerability.

We determine the time after the first case in each country based on the weight calculation from the vulnerability. In the first stage, the daily confirmed cases number is determined by a random number based on the historical data distribution of the daily confirmed cases number and vulnerability in each country. In other words, if a country has a high number of confirmed cases during the first stage, it means that the country either lacks adequate medical conditions and epidemic control measures or is highly vulnerable to China. It is very likely that there will be a higher chance of a confirmed case in the future. In our results section, we estimate and calibrate the vulnerability and the number of confirmed cases for each country.

According to the available data and examples of local cluster infections in Singapore, >40 cases are confirmed, which means that the potential for cluster infections is quite high. After entering community cluster infections, the influence of each country’s own populace is greater than that of immigrants. Therefore, it is defined as entering the second stage of simulation after the cumulative case number is >40 cases.

#### Second stage: Localized outbreaks

It is assumed that in the second stage—after entering the local cluster infection— the pattern change in the number of cases will be similar to that of Hubei Province or other provinces in China. Since this study assumes that future information is clearer than when China started its outbreak, all countries have already prepared to prevent epidemic outbreaks. Therefore, a more conservative method is used to carry out Monte Carlo simulation using data from provinces other than Hubei. Generate 1,000 simulation data of future case growth and extract three scenarios of high (quantile = 0.95), medium (quantile = 0.5), and low (quantile = 0.05) infection from the simulation results while retaining the potential for changes in the number of cases in Hubei as the worst future scenario (Extreme Scenario). Both a sigmoid function and polynomial regression are used to extend the simulation time for different assumed second scenarios, A sigmoid function is used to assume that the epidemic in China has almost reached its peak as a more conservative scenario; polynomial regression is used for the assumption that the epidemic in China will continue and shows no signs of easing (Table 1).

**Table 1.** The setting of simulation scenarios.

| First Scenarios | Second Scenarios |  |
| --- | --- | --- |
|  | 1. Conservative | 2. Severe |
| **A. Extreme** | A1: Extreme–Conservative | A2: Extreme–Severe |
| **B. High (quantile = 0.95)** | B1: High–Conservative | B2: High–Severe |
| **C. Medium (quantile = 0.5)** | C1: Medium–Conservative | C2: Medium–Severe |
| **D. Low (quantile = 0.05)** | D1: Low–Conservative | D2: Low–Severe |

Monte Carlo simulation is a very suitable risk-assessment method; it can perform simulation based on limited data and different scenarios to understand the possible risks since computers can easily simulate a huge number of experimental trials that have random outcomes and uncertainty (Papadopoulos & Yeung, 2001). The elements of the Monte Carlo simulation method including the expectation *E*_*π*_{*U*(*X*)}, which is with respect to the probability density π, the response function *U(x)*, and random draws *X = x(j)* from the target distribution π (Jørgensen, 2000).

### c. Risk assessment

Based on the Assessment Report published by the IPCC (Field, Barros, Stocker, & Dahe, 2012; Pachauri et al., 2014; Parry et al., 2007). The risk assessment is based on the results of the interaction of various components: hazard, vulnerability, and exposure. In other words, the risk of adverse effects from the impact of extreme events is defined as a function of the vulnerability, exposure, and hazard. Here, extreme events would be outbreaks of COVID-19 in global countries. Among them, we adapt the definition of vulnerability from IPCC; vulnerability is the degree to which a system is susceptible to and unable to cope with the adverse effects of climate change, including climate variability and extremes (Parry et al., 2007).

Therefore, according to the most commonly used approach to risk definition, the risk is defined here as:

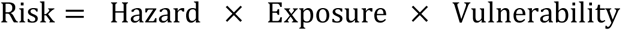

The infectivity and mortality of COVID-19 under different climatic conditions remain unknown at present, so it is assumed that the infectivity of COVID-19 and the hazard to the human body are the same in all countries. The degree of hazard in each country becomes the estimated number of cases within the country; the greater the number of cases, the greater the potential hazard. Exposure is each country’s population density. We assume that countries with higher population densities are more likely to be exposed to COVID-19. Under the condition that local cluster infection has occurred in the second stage, the higher the population density, the more likely a serious outbreak and thus higher overall risk. Vulnerability is defined as the ratio of connectivity to the Health Care Index calculated in this study, as previously described. At present, only countries’ vulnerability to China is considered. In the future, it can be summed according to whichever countries have experienced large-scale outbreaks based on the superposition principle.

## Result

### a. Calibration of China’s confirmed case number

Since the local medical system in Hubei Province may be insufficient to support the huge number of patients, the official diagnoses may be an underestimate. Therefore, we first collected information on the evacuation flights from different countries at different times to calculate the proportion of confirmed cases and used this to estimate the infection rate in Wuhan based on the concept of statistical sampling, as shown in Table 2.

**Table 2.** The confirmed cases and number of evacuation flights (source: Collected from the international press)

| Date | Country | Number<br>of people<br>on flights | Confirmed<br>cases | Suspected<br>cases | Confirmed<br>cases after<br>quarantine | Proportion of<br>confirmed<br>cases (%) | Remark |
| --- | --- | --- | --- | --- | --- | --- | --- |
| 2020/1/29 | Japan | 206 | 3 |  | 1 | 1.94 | Two asymptomatic cases |
| 2020/1/29 | United States | 195 | 0 |  |  | 0.00 |  |
| 2020/1/30 | Japan | 210 | 2 |  |  | 0.95 |  |
| 2020/1/31 | Japan | 149 | 3 |  | 1 | 2.68 |  |
| 2020/1/31 | France | 180 | 0 | 1 |  | 0.00 |  |
| 2020/2/1 | Germany | 124 | 2 |  |  | 1.61 |  |
| 2020/2/2 | France | 254 | 0 | 36 |  | 0.00 | The test for suspected cases are all negative |
| 2020/2/2 | Indonesia | 237 | 0 |  |  | 0.00 |  |
| 2020/2/3 | Malaysia | 141 | 2 |  |  | 1.42 |  |
| 2020/2/3 | Australia | 243 | 0 |  |  | 0.00 |  |
| 2020/2/3 | Vietnam | 194 | 0 |  |  | 0.00 |  |
| 2020/2/4 | Taiwan | 247 | 1 | 3 |  | 0.40 | Confirmed patients and fever patients are not allowed to board aircraft |
| 2020/2/4 | Thailand | 138 | 0 |  | 1 | 0.72 | Two of the original 141 people did not board the plane because of fever |
| 2020/2/5 | United States | 350 | 0 |  |  | 0.00 |  |
| 2020/2/5 | Russia | 80 | 0 |  |  | 0.00 |  |
| 2020/2/6 | United States | 410 | 0 |  | 1 | 0.00 |  |
| 2020/2/7 | Japan | 198 | 1 | 4 |  | 0.51 |  |
| **SUM<sub>1</sub>** |  | **3556** | **14** | **44** | **4** | **0.51** |  |
| **SUM<sub>2</sub>**<br>(Japan, Germany, Malaysia) |  | **1028** | **13** | **4** | **2** | **1.46** |  |

Sample 1 was calculated based on the average of all cases in the countries collected in Table 2. The proportion P_1_ = 0.51%, with 95% confidence interval [0.32%, 0.80%]. There are nine million current residents in Wuhan as announced by the press conference held by the Hubei Provincial Government of China on the evening of 1/26; of these, approximately 45,900 people with 95% confidence interval [28,800, 72,000] may be infected. As of the official announcement of 2020/2/8, the number of confirmed cases in Hubei has reached 27,100 people, and the number of confirmed cases in China has reached 34,546 people. This estimation sample is insufficient to show the number of hidden cases.

According to the news press and public information, avoiding onboard infection or the triggering of more domestic infections, many countries have screened evacuating people, such as those who have a fever or with confirmed infections, to prevent them from boarding planes. Therefore, this sampling will be underestimated.

Sample 2 only collected cases of Japanese, German, and Malaysian evacuation charter flights as these may be more relevant for sampling. In other words, the worst scenario is taken here. The proportion P_2_ = 1.46%, with 95% confidence interval [0.89%, 2.39%]. Based on the nine million people living in Wuhan, the average number of people infected during early February was about 131,400 with 95% confidence interval [80,100, 215,100].

### b. Vulnerability

Vulnerability to the COVID-19 outbreak from China is calculated for 63 countries around the world, as shown in Figure 2. Table 3 shows the first 20 countries’ vulnerability. Taiwan has the highest vulnerability to novel coronavirus outbreak from China, or broadly speaking to various human infectious diseases from China, followed by South Korea, Hong Kong, and Japan. These countries are the most vulnerable, followed by Thailand, the United States, Russia, Macau, and Singapore. The top-20 countries listed in Table 3 are considered relatively vulnerable.

**Table 3.** The first 20 countries’ vulnerability to the COVID-19 outbreak from China (without China).

| Rank | Country | Vulnerability |
| --- | --- | --- |
| 1 | Taiwan | 1.000 |
| 2 | South Korea | 0.848 |
| 3 | Hong Kong | 0.809 |
| 4 | Japan | 0.791 |
| 5 | Thailand | 0.426 |
| 6 | United States | 0.387 |
| 7 | Russia | 0.373 |
| 8 | Macau | 0.326 |
| 9 | Singapore | 0.287 |
| 10 | Malaysia | 0.194 |
| 11 | Vietnam | 0.182 |
| 12 | Cambodia | 0.120 |
| 13 | Germany | 0.118 |
| 14 | United Arab Emirates | 0.101 |
| 15 | Australia | 0.099 |
| 16 | Indonesia | 0.097 |
| 17 | Burma | 0.091 |
| 18 | Philippines | 0.087 |
| 19 | Netherlands | 0.078 |
| 20 | Canada | 0.075 |

**Figure 2.**
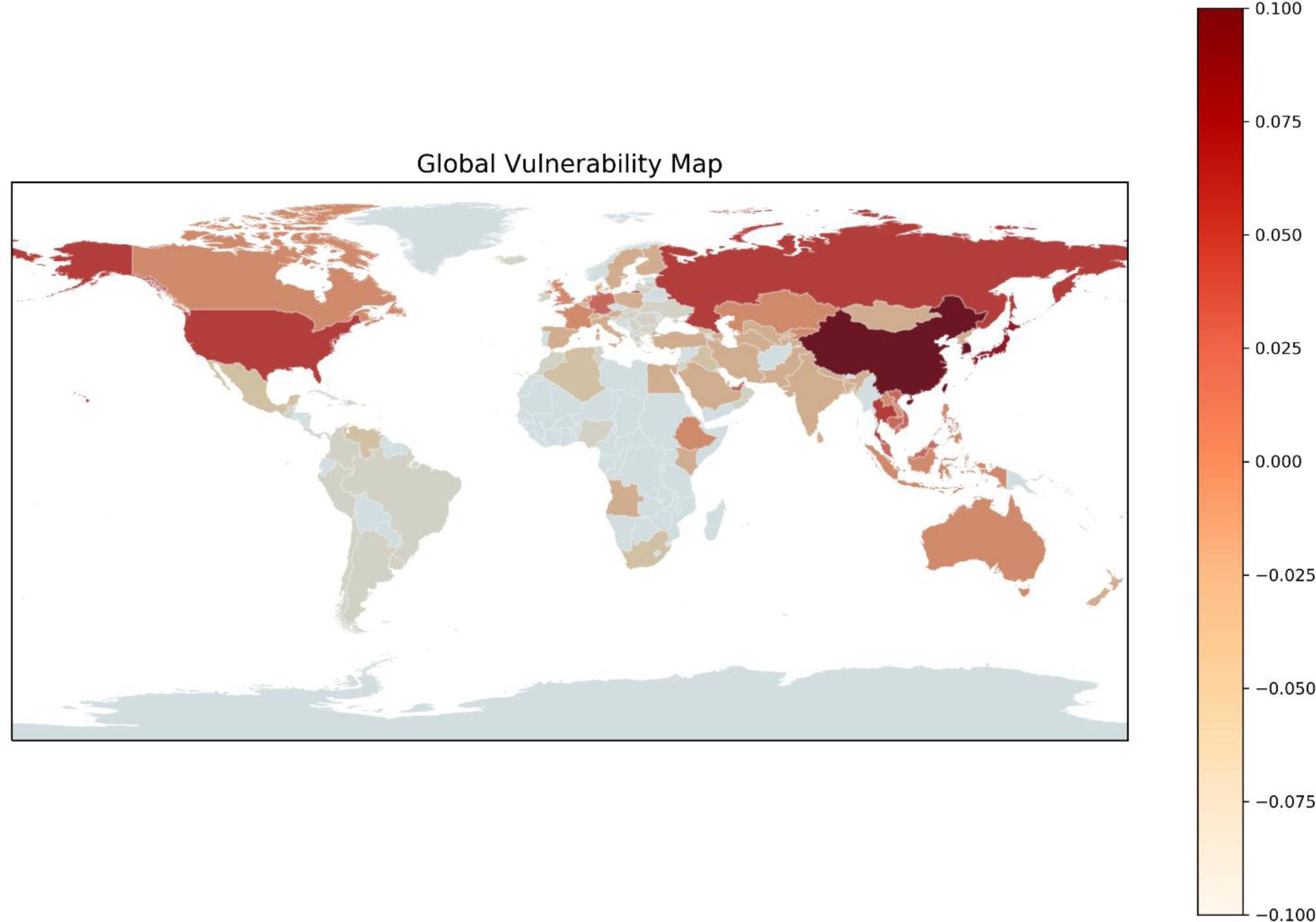
Global vulnerability map for the COVID-19 outbreak from China

Without effective anti-epidemic measures in these countries, cases of overseas imports from China may soon occur. Figure 3 observes the relationship between each country’s vulnerability and days taken for the first imported case to occur; they show a very high exponential decreasing relationship. This result proves our argument and confirms that the vulnerability index proposed by this study is very informative and can well express the possibility and number of cases that may happen in the first stage for each country. Although different countries’ immediate epidemic prevention policies may affect this; overall, the more vulnerable a country, the sooner its first overseas import case from China would occur. Meanwhile, the lower the vulnerability, the later such a case would occur.

**Figure 3.**
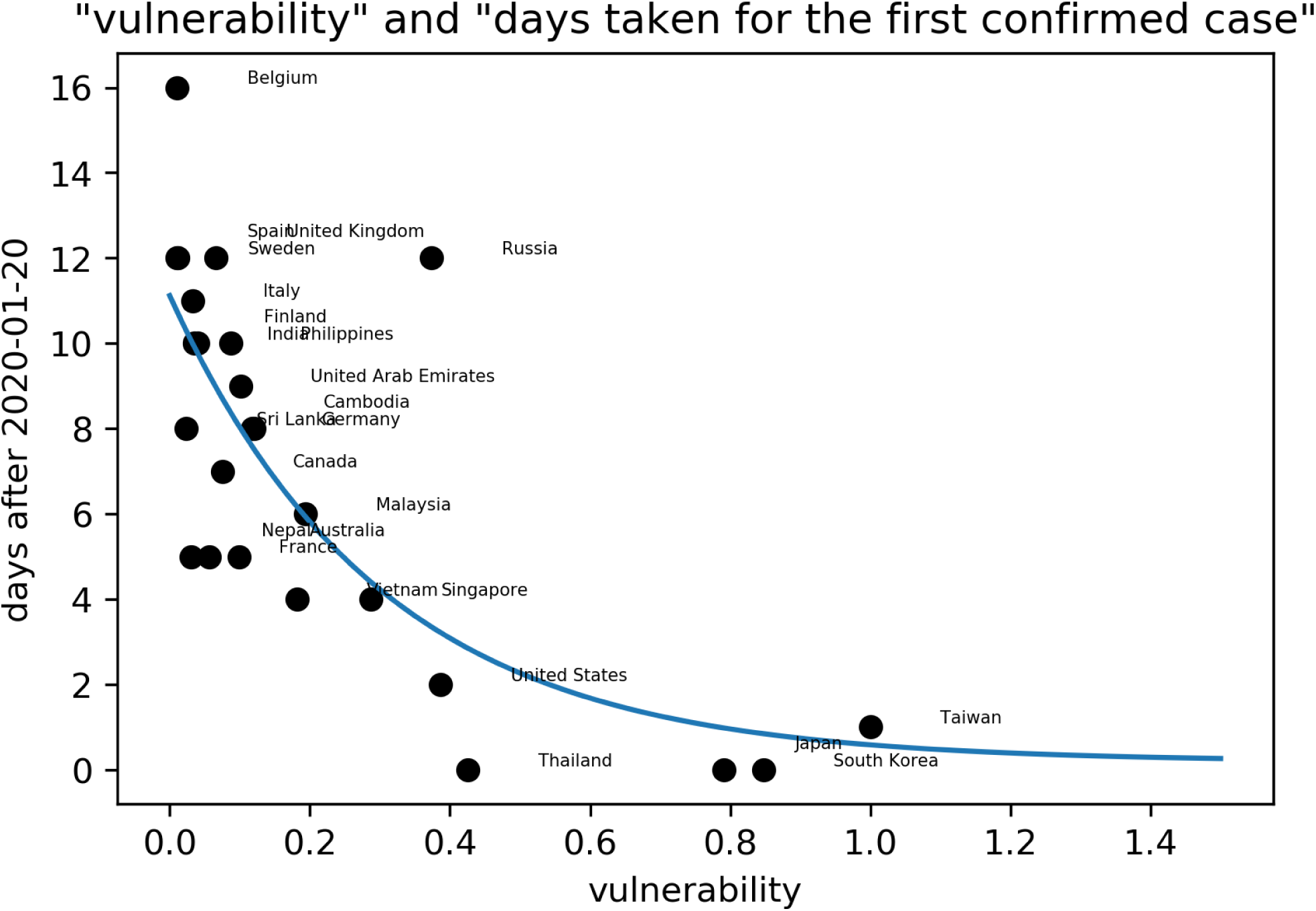
The relationship between each country’s vulnerability and how many days it took before the first imported case occurred. The fitted regression function is *y = 10*.*93exp(−3*.*32x) + 0*.*19*

In addition, we can observe the relationship between vulnerability and the cumulative number of cases in each country as shown in Figure 4; this shows a clear linear relationship. This is a dynamic process that changes daily with the number of cumulative cases. Since the beginning of the outbreak in January 2020, various countries’ governments have implemented related measures for epidemic prevention such as reducing flights and stopping visas so that many countries with high vulnerability can maintain a small number of cases. The countries below this regression line—including Taiwan, Korea Republic, and Russia—can be regarded as countries with excellent initial epidemic prevention work and Japan and the United States have performed fairly well (unfortunately, Japan starting finding outbreak community infections after this study was completed), whereas Singapore and Thailand are far above the regression line with higher cases and may have higher risk in the future.

**Figure 4.**
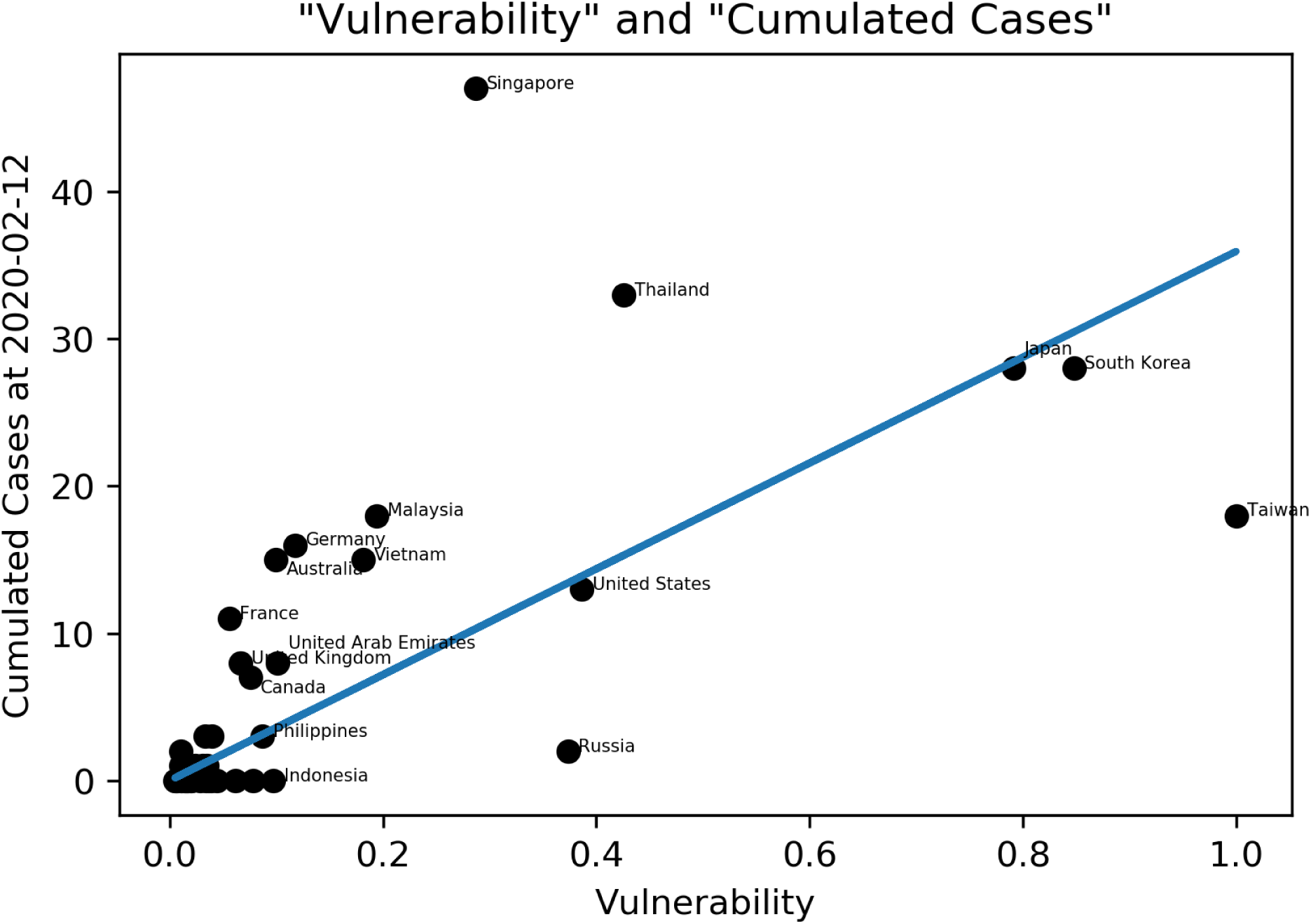
The relationship between vulnerability and the cumulative number of cases in each country. The coefficient of the OLS Regression equation is 35.94 with R^2^ = 0.587 and 95% C.I = [26.896, 44.994].

### c. Simulation result and dynamic risk analysis

Figure 5 is the time series of the 1,000 countries’ simulated case numbers after the second stage outbreak in the future using Monte Carlo simulation. Figure 6 is the simulated number of potential cases under four different scenarios: high, medium, and low, and the extreme scenarios of Hubei Province. After combining the simulation results of the countries’ first and second stages around the world using the data fusion algorithm, the simulated cases in different future scenarios are generated as shown in Figures 7 and 8.

**Figure 5.**
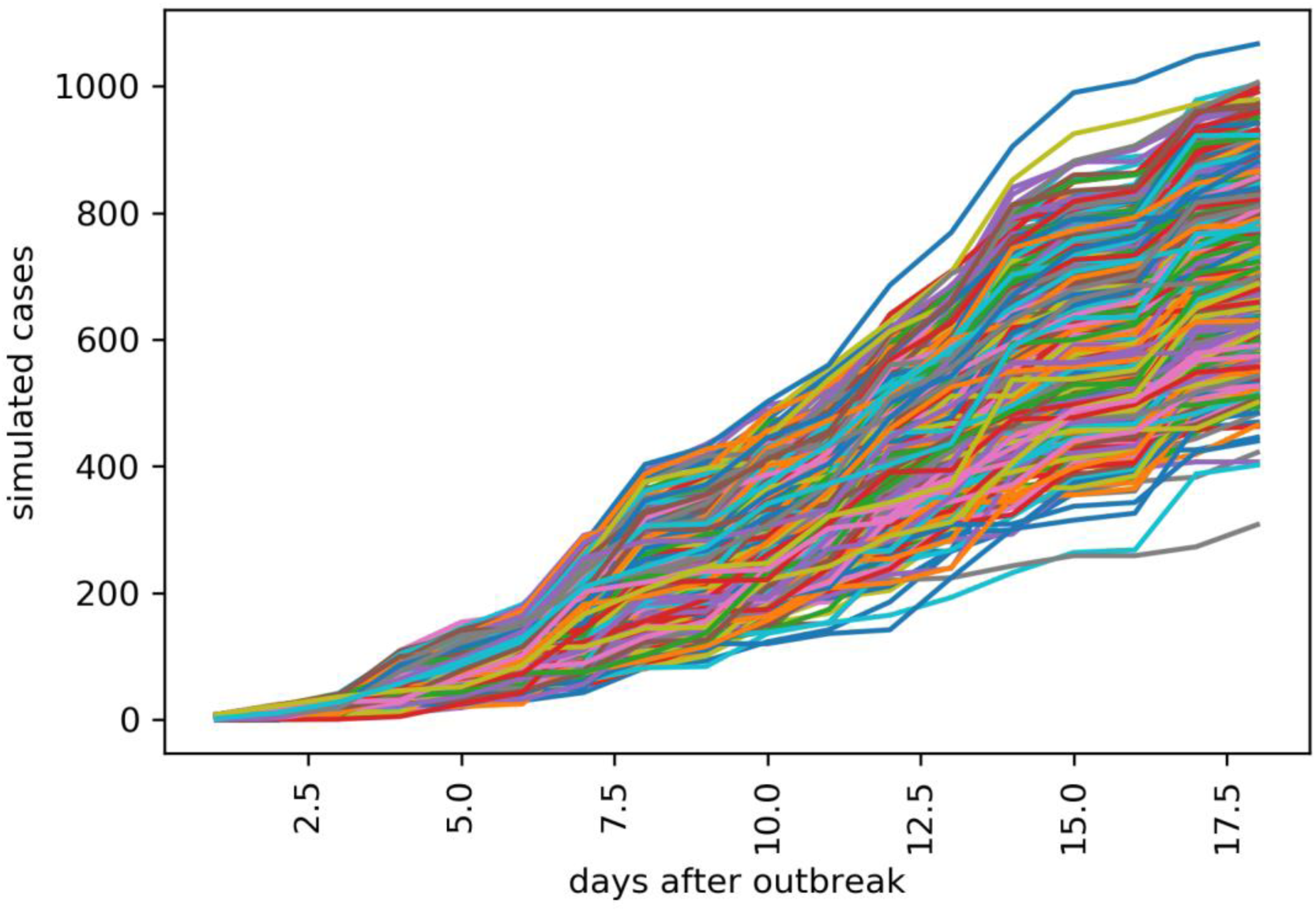
Time series of the 1,000 countries simulated case numbers after the second-stage outbreak in the future using Monte Carlo simulation.

**Figure 6.**
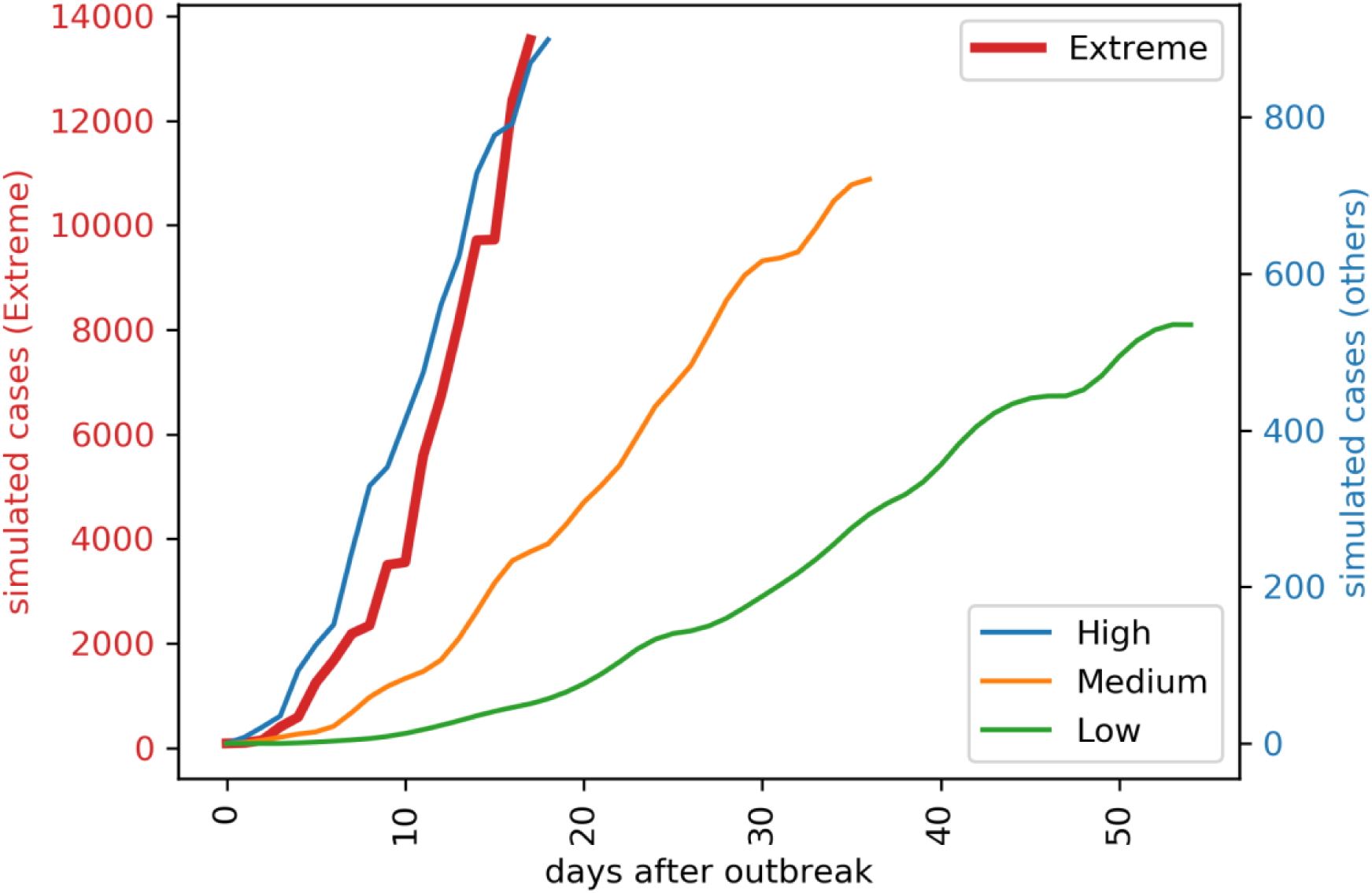
The simulated number of potential cases in four different scenarios: high, medium, low, and the extreme scenarios of Hubei Province

**Figure 7.**
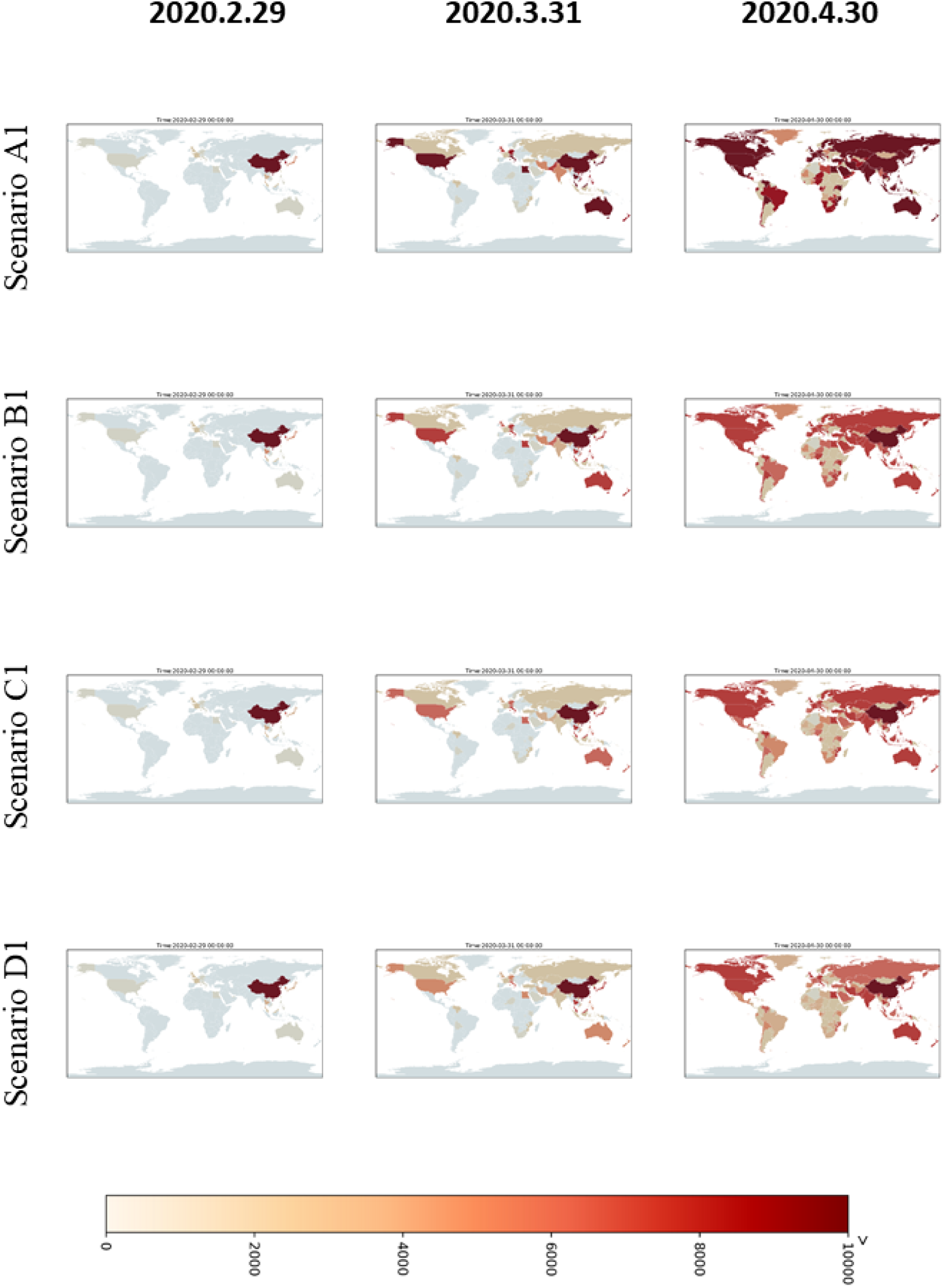
The simulated cumulative case maps for conservative scenarios of A1, B1, C1, and D1 in different time-slices.

**Figure 8.**
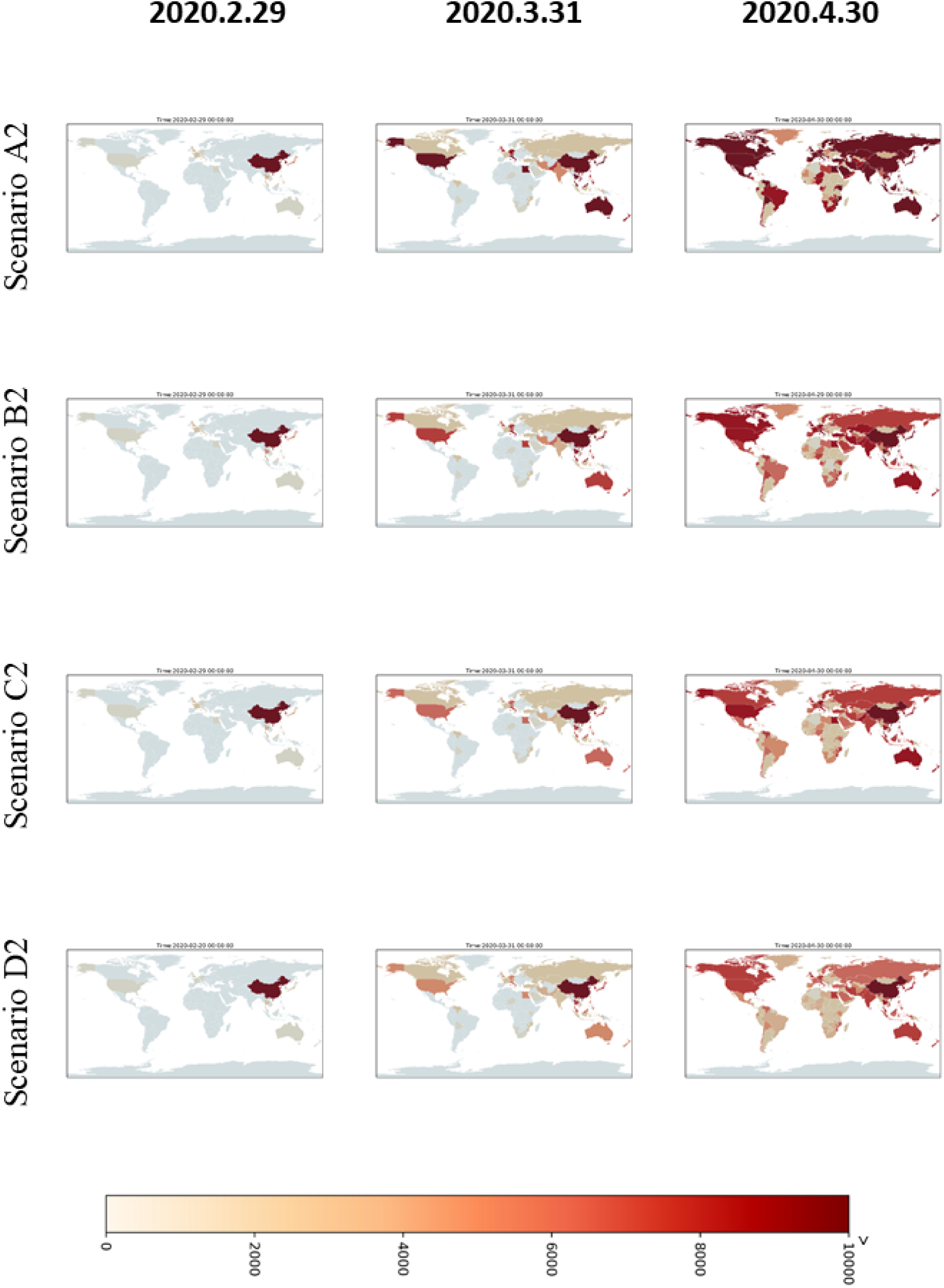
The simulated cumulative case maps for severe scenarios of A2, B2, C2, and D2 in different time-slices.

According to the spatiotemporal distribution results of different scenario simulations, the conservative scenarios (Figure 7) assume that China has entered the start of its peak period and the number of outbreak cases has started to decline. Therefore, the results of simulations applied in other countries show that the cumulated case number increases rapidly at the start for some countries, particularly for countries with higher vulnerability. After one month, the epidemic was controlled and the number of cases in each country no longer increased significantly. Meanwhile, considering the extreme, high, medium, and low simulation levels of the epidemic scenarios in various countries, it can also be seen that after a period of time, namely in April, the case number differences between the scenarios will be clearly comparable. A1 extreme is the highest scenario, followed by B1, C1, and D1. Meanwhile, severe scenarios (Figure 8) are based on the trend that COVID-19 may be like traditional influenza for a global pandemic; that is, cases will continue to occur, which may lead to >10,000 cases in some countries. However, the general spatial distribution of case numbers is similar to conservative scenarios. This scenario is still the same as that with the highest case numbers in the A2 extreme scenario, followed by B2, C2, and D2.

Then, the risk calculation method combines hazard with vulnerability and population density to calculate changes over time in the different scenarios of different countries around the world over time to generate the COVID-19 dynamic risk map, as shown in Figure 9. Here, we show some representative time slices only with scenario C2. For the full time period 2020/1/21–2020/5/31 in the GIF animation format, please refer to Supplementary Materials. Due to each country’s different vulnerability and population density, the spatial distribution will differ slightly from the hazard maps show in Figures 7 and 8. Some countries have higher risks because of their higher vulnerability or population density. In particular, Asian countries near China have higher risk, particularly East and Southeast Asia. Some other countries have gradually increased their risks over time. For example, countries in Europe, Africa, and South America started increasing their risks from March to April.

**Figure 9.**
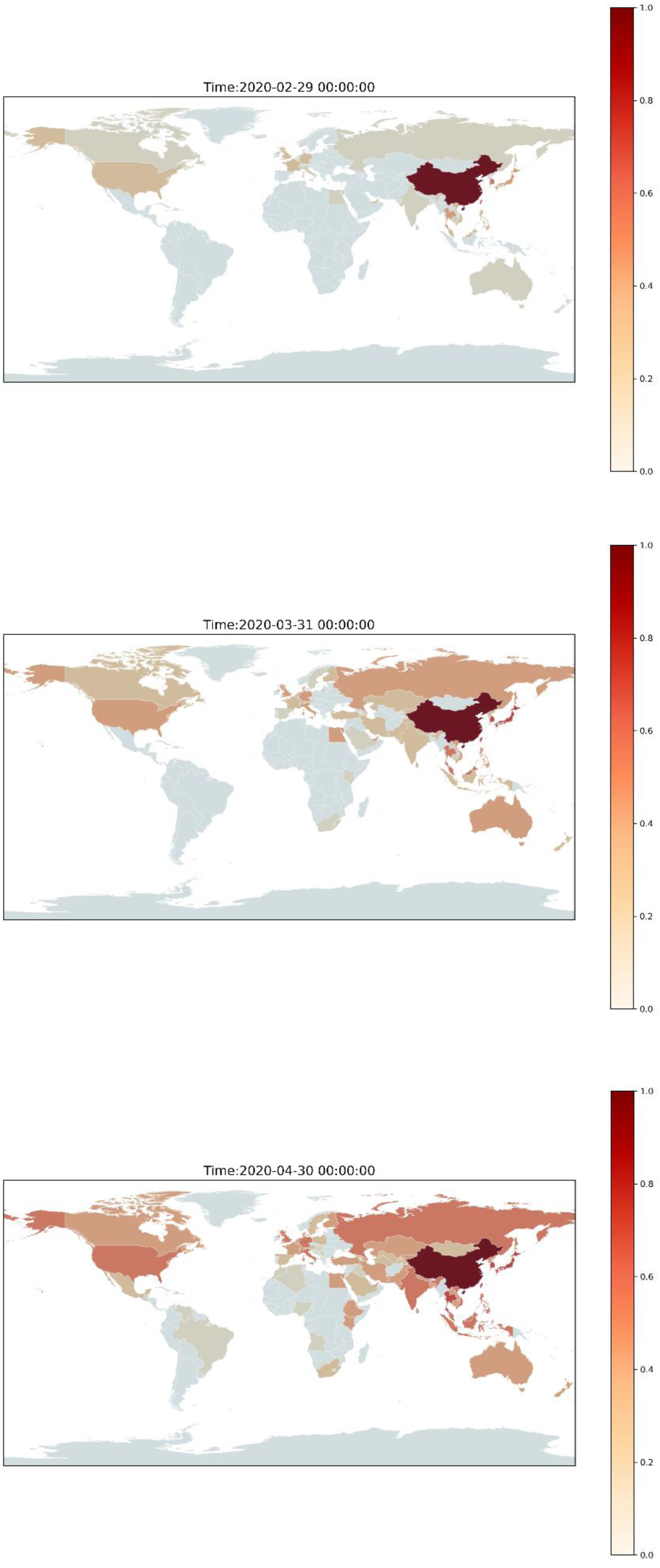
A dynamic risk map for scenario C2 at different time slices (a) 2020/2/29, (b) 2020/3/31, and (c) 2020/4/30.

## Discussion

This study proposes a framework for the dynamic risk analysis of novel coronavirus SARS-CoV-2-infected pneumonia disease (COVID-19), attempts to understand the impact of vulnerability, and uses this information to simulate the development of the number of potential cases in different countries in the future. In addition, this study compares and quantifies the initial epidemic prevention capabilities and various countries’ management strategies. At present, this study focuses on the spread of risk in space and time, which can be used as early-stage control when a novel infectious disease breaks out. In the future, it should be possible to simulate the changes in the number of recoveries or deaths at the same time to better understand the future risk reduction process. Based on this framework, we can perform the next stage of simulation, continuously modify the risk map, and dynamically update the analysis results. However, there is currently no complete understanding of the characteristics of COVID-19 around the world. There is still a great deal of uncertainty regarding when the epidemic will reach its peak and it is unclear whether recovered patients are still infectious.

Although the outbreak occurred in China, the epidemic is still in its initial stage for the whole world. This study is a preliminary estimation and it can be assumed that COVID-19 is not controlled by climatic factors or effective drugs. Moreover, many cases with mild symptoms or even asymptomatic cases have appeared clinically, which has made epidemic prevention and modeling very difficult. According to this study’s simulation results, if there is no specific medicine, it will likely form a global pandemic.

Although in this study, we collected the proportion of evacuation charter infection cases in various countries and estimated that the total number of cases in Hubei Province is much larger than the actual official case number, we still adopt a more conservative method that uses the official case number for risk assessment. This is based on the assumption that countries are well-prepared for epidemic prevention. We believe that countries have been prepared and have relevant experience for reference earlier than the period when the Wuhan outbreak started. In the next stage, based on the research framework, we can further superimpose the next outbreak on the global impact such as the localized infection that happened in Singapore, and so on.

The other results of many statistical models will be greatly affected by adjusting the parameters. At the start of an unknown infectious disease outbreak, it is impossible to understand the actual parameters. For example, we have been unable to determine the basic reproductive number, serial interval, etc. These will affect the model’s prediction results and create difficulties in model prediction. In fact, we have also tried to use a data-driven approach to make estimations using machine-learning or deep-learning methods such as Long Short-Term Memory (LSTM) and the Random Forest method. However, the preliminary results are poor due to insufficient data and cannot be effectively used for supervised learning.

After completing this study, it was also found that localized outbreaks in Japan and Singapore had taken the lead, consistent with the vulnerability analysis and risk analysis results in this study. Both Singapore and Japan were among the countries with the highest risk in the simulation results. The first case also occurred in Egypt (risks signal exist in the Egyptian part of the map as of February 20, 2020 in each scenario of Figures 7 and 8). This is consistent with the estimation results in this study.

## Data Availability

The authors confirm that the data supporting the findings of this study are available within the article.

## Acknowledgments

We are grateful to the funding support from the Ministry of Science and Technology (MOST) in Taiwan, project no. MOST 108-2636-E-008-004 (Young Scholar Fellowship Program). We would also like to thank the Python programing language and its modules as a powerful tool in our data analysis and the raw data providers described in the article.

## Author Affiliations

YCL is from the Department of Civil Engineering, Research Center for Hazard Mitigation and Prevention, National Central University, Taiwan. The other authors are from the Department of Civil Engineering, National Central University, Taiwan

## Contributors

YCL designed the experiments, YCL, CYL, and YTL collected data, YCL and WJC analyzed the data, and YCL interpreted the results and wrote the manuscript.

## Supplementary Materials

SP 1. A GIF animation of the global dynamic risk map for COVID-19 under the scenarios of C2.

